# School performance and mortality in young adulthood: a register-based population study

**DOI:** 10.1101/2023.03.15.23287297

**Authors:** Bjørn-Atle Reme, Fartein Ask Torvik

## Abstract

**Background:** The social gradient in mortality among young adults is well-documented, but poorly understood. This study contrasts the role of parental income and education versus own school performance as predictors of early death.

**Methods:** We estimated the prospective association between grade point average from lower secondary education and mortality between age 16 and 30 for the Norwegian birth cohorts 1985-2002 (N = 986,573). This association was compared to the associations between measures of family socioeconomic status and young adult mortality in the offspring. We also estimated hazard ratios in models including both school performance, parental income, and education. Sibling comparison analyses were also estimated to assess the importance of within-family variation. Last, we used the cause of death register to estimate the hazard ratio of different causes of death among individuals with poor school performance.

**Findings:** During the observational period, we observed 1300 deaths in the lowest quartile of school performance (0·06%) and 413 deaths in the highest quartile of school performance (0·02%). The risk of early death among boys in the lowest quartile of school performance compared to the highest quartile was larger for boys [HR = 3·68, 95% CI 3·54-3·81] than for girls [HR = 2·82, 95% CI 2·62-3·01]. The corresponding hazard ratios across parental income quartiles were 1·79 [95% CI 1·67-1·1·91] for boys, and 1·63 [95% CI 1·43-1·82] for girls. Across parental education level, the hazard ratio was 2·03 [95% CI 1·85-2·21] for boys, and 1·64 [95% CI 1·35-1·93] for girls. When jointly including school performance, parental income and education in the same model, parental income and education were insignificant, while the association school performance remained strong: HR = 3·57 [95% CI 3·44-3·71] for boys, and 2·98 [95% CI 2·78-3·18] for girls. With regards to causes of death, the highest hazard ratio among those in the lowest quartile of school performance was for drug-related poisoning, with 6·47 [95% CI 2·78-3·18] for boys and 7·3 [95% CI 4·93-10·80] for girls. The results were consistent in sibling comparison analyses.

**Interpretation:** School performance is a substantially stronger predictor of early death than common measures of socioeconomic background. School performance absorbs the social gradient in early death.

**Funding:** This work was supported by the Research Council of Norway (grant number 273659). This work was partly supported by the Research Council of Norway through its Centers of Excellence funding scheme (grant number 262700).

## Introduction

Some die young. The first step in preventing these tragedies it to know who is at risk. Although studies consistently find higher mortality risk among individuals with lower socioeconomic status, ^1-7^ less is known about the mechanisms underlying inequalities in young adult mortality. Clearly, socioeconomic status is not a direct cause of death. However, it is an indicator of deprivation, differences in resources, health behaviors, or other causes that starts a cascade of events that eventually influence the risk of dying.^8, 9^ To better understand, and possibly prevent, early adult death, it is therefore critical to develop knowledge about factors can predict death in early adulthood. Previous studies have documented the important role of family background characteristics, such parental education or income.^10, 11^ Other studies point to the importance of educational attainment for understanding mortality.^4, 12-16^ While the gradient in mortality across family background and educational attainment is well established, there are no large-scale investigations of their relative importance. Moreover, the role of indicators observable earlier in life, such as school performance at age 16, is largely unexplored.

In this study we contrast the role of socioeconomic background with an individual’s own school performance as predictors of early death. Our aim was to advance the existing literature by (1) estimating the association between school performance, defined as Grade Point Average (GPA) at age 16 and death in young adulthood, (2) examine the role of parental education and income for early death, (3) examine the relative strength of GPA and family background in explaining early death, and (4) estimate how the risk of cause-specific early death varied across school performance.

## Methods

### Study design and participants

The study was based on population-wide administrative data from 4 Norwegian national registries: The Population Register, The National Education Database, the National Registry for Personal Taxpayers, and the Cause of Death Registry. The study was approved, and participant consent was waived by the Regional Committee for Medical and Health Research Ethics South-East Norway (REK, approval 2018/434).

This population-based cohort study included all individuals born in Norway between 1985 and 2002, who resided in the country during their lower secondary education (age 13-16) and received a GPA (N = 986,573). We excluded individuals where both parents missed income or education data. Figure S.1 in the Supplementary material provides a graphical representation of the sample selection procedure.

### Measures

#### School performance

The National Education Database contains complete grade records from lower secondary education for graduation years 2001 to 2018. Lower secondary education, 8^th^ to 10^th^ grade, is compulsory for Norwegian citizens. The GPA score is calculated as the average of all courses and exams completed during lower secondary education. Grades range from 1 to 6, where 6 is best. It is not possible to fail lower secondary school, hence there is no sample selection in GPA. However, a small fraction (4%) does not receive a GPA, typically due to a high level of absence where the teacher cannot grade the student’s performance. These students are still able to proceed to tertiary education on special terms. In Norway and many other counties, GPA is used for ranking students for placement in upper secondary education. In the analysis we divided the individuals into gender and birth cohort stratified GPA quartiles. Throughout the analysis, the quartiles are referred to as Q1 (low), Q2, Q3 and Q4 (high).

#### Measures of parental socioeconomic status

Socioeconomic status was measured in two ways, by parental education level and household income. Parental education level was retrieved from the National Education Database using the highest level of completed education among the individual’s parents at age 16. Household income quartiles were estimated by summing parental taxable incomes in the year the child was 10, and quartiles then created within each birth cohort of the child, i.e., relative to other 10-year-old children in the given year.

#### Causes of death

The year and cause of death were obtained from the Cause of Death Registry. It contains information on 96·3% of all deaths in the relevant study period, 2002 to 2018. Underlying causes of death were originally classified according to the International Classification of Diseases, Tenth Revision (ICD-10). In the data available for this study the causes of death were classified according to the European Shortlist Causes of Death version 1998 (see Supplementary Material Table S.2 for a complete list, and correspondence with ICD-10).^17^

### Statistical analysis

We estimated the sex-specific young adult mortality risk, between ages 16 and 30, using Cox regressions. The main analyses included four different models in terms of included covariates, estimated separately for each sex: (i) school performance (binary indicators GPA quartile), (ii) household income (binary indicators of family income quartiles), (iii) parental education at age 16 (binary indicators of highest parental educational attainment), and (iv) all these, (i)-(iii), simultaneously. From these models we report the sex-specific predicted number of young adult deaths per 10 000 and the young adult mortality relative risk ratios (RRs), with the highest level within each type of covariate as the reference category (Q4 of school performance, Q4 of family income, and master/PhD-level parental university education).

To further examine the robustness of our findings, and adjust for family-specific factors that may affect both GPA and the young adult mortality risk, we estimated sibling models, i.e., utilizing only intra-family variation (frailty at the mother level).^18^ In order to make estimates directly comparable, the benchmark model and the sibling models were restricted to including only individuals with at least one other sibling with a GPA record (N = 740,184).

In the second part of the analysis, we utilized the cause of death register to examine how registered causes of early adult deaths differed across school performance. We do so by first estimating the number of deaths from different causes within each quartile of school performance, then by estimating hazard ratios for different causes of death for the lowest quartile of school performance relative the other quartiles.

## Results

Our data covered birth cohorts 1985-2002 and was right censored in 2018. During the observational period, 3083 (0·3%) individuals died (1300 (0·06%) in Q1 and 413 (0·02%) in Q4). Since our data is right-censored, these numbers are lower than the full young adult mortality risk between age 16 and 30. In the remaining analysis we therefore estimate the young adult risks and likelihoods using Cox regressions.

### School performance and young adult mortality

Figure 1 shows the estimated likelihood of death between ages 16 and 30 for each quartile of school performance, separately for each gender. The estimates were derived from a Cox regression (see Table S.1 in the Supplementary material for corresponding table). Relative to the highest quartile of school performance, the hazard ratio in the lowest quartile was higher among boys (HR = 3·68; 95% CI 3·54-3·81) than girls (HR = 2·82; 95% CI 2·62-3·01) (see Table S.4 in the Supplementary material for a table of the hazard ratios).

**Figure 1:**
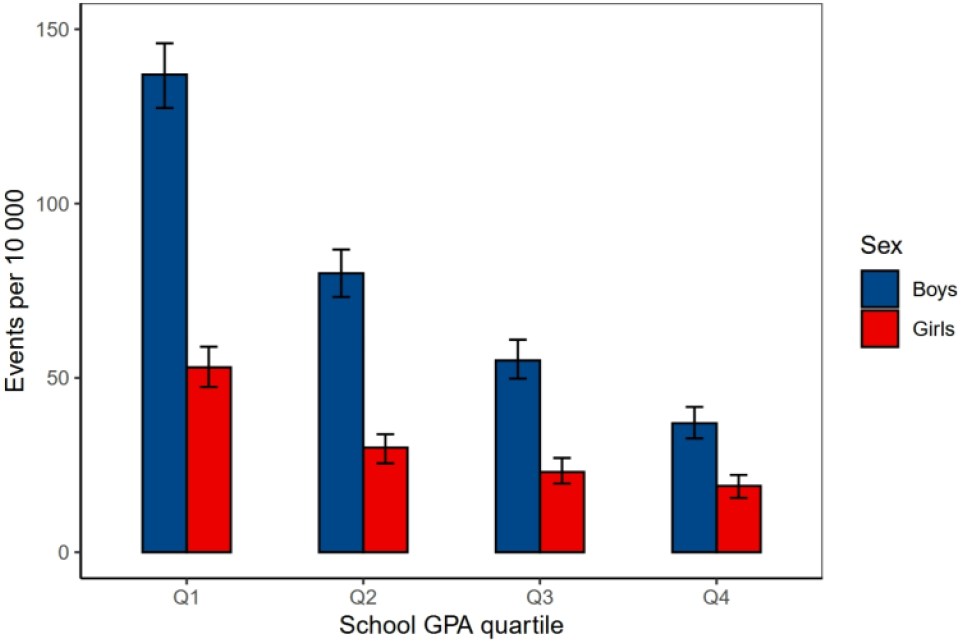
Estimated number of young adult deaths across sex and quartiles of school performance. Predicted number of young adult deaths between age 16 and 30 from a Cox regression, estimated separately for each sex and using dummy variables for each quartile of school performance. See Table S.1 in the Supplementary material for a corresponding table.

### Parental socioeconomic status and young adult mortality

Figure 2 shows the predicted likelihood of death between ages 16 and 30 for each quartile of household income (panel A) and parental education level (panel B), separately by sex (see Table S.2 and S.3 in the Supplementary material for corresponding tables). Relative to the boys from the highest quartile of household income, the hazard ratio among boys from the lowest quartile was 1·79 [95% CI 1·67-1·91]. The corresponding ratio was 1·63 [95% CI 1·43-1·82] for girls. The hazard ratios across parental education level, from lowest to highest levels of education was 2·03 [95% CI 1·85-2·21] for boys, and 1·64 [95% CI 1·35-1·93] for girls (see Table S.4 for a table of hazard ratios).

**Figure 2:**
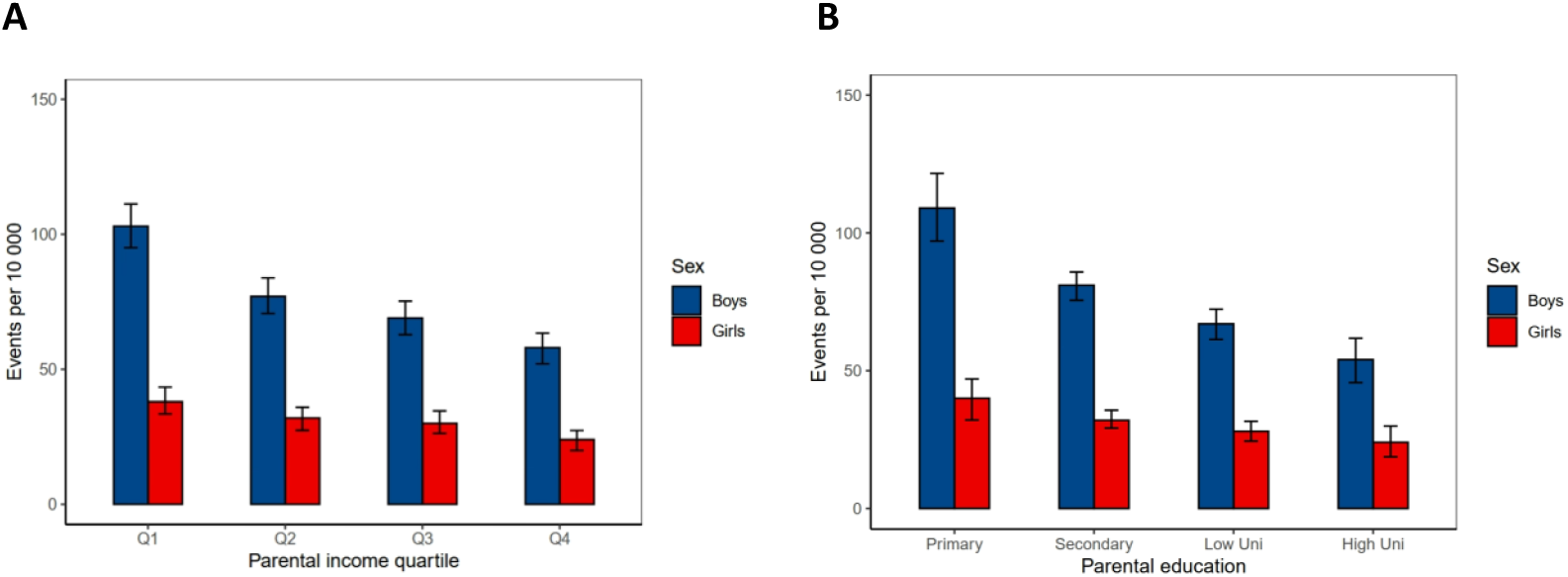
Estimated number of young adult deaths across sex, parental income and parental education level. Predicted number of young adult deaths between age 16 and 30 from a Cox regression, estimated separately for each sex and using dummy variables for each quartile of parental income (A) and highest level of parental education (B). Panel A had N_Q1_=223,642; N_Q2_=253,079; N_Q3_ =256,366,642; N_Q4_=253,486. Note that the slight imbalance in number of observations was due to quartiles being created based on all children in a given birth cohort, also including those with missing GPA. Panel B had N_Primary_ = 90,758; N_Secondary_=427,823; N_LowUni_ =340,020; N_HighUni_ = 127,972. See Table S.2 and S.3 in the Supplementary material for corresponding tables.

### School performance and parental socioeconomic status combined

Figure 3 shows the proportional hazards for death between ages 16 and 30 estimated using a Cox regression including school performance, parental education and parental income as explanatory variables (see Table S.4 in the Supplementary material for a table). The model was estimated separately for each gender. While the coefficients for the different levels of household income and parental education are close to 1, the coefficients with respect to school performance are not. Relative to the boys in the highest quartile of school performance, the hazard ratio among boys from the lowest quartile was 3·57 [95% CI 3·44-3·71], and 2·98 [95% CI 2·78-3·18] for girls.

**Figure 3:**
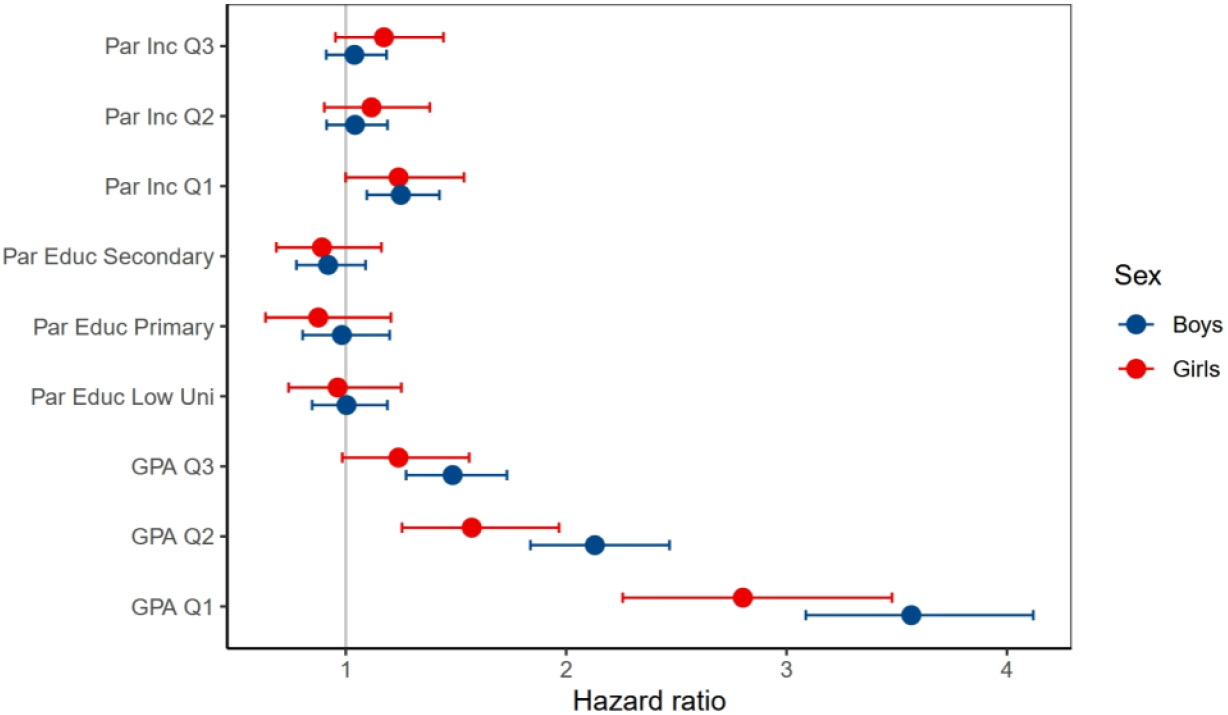
Estimated number of young adult deaths across sex, school performance, parental income and parental education. Predicted number of young adult deaths between age 16 and 30 from a Cox regression, estimated separately for each sex and using dummy variables for each quartile of school performance, quartile of parental income, and highest level of parental education. See table S.4 in the Supplementary material for a corresponding table.

### Sibling comparions

The benchmark model for boys - same specificaction as above (Figure 3), but including only individuals with at least one sibling with a GPA record - the hazard ratio was 3·41[95% CI 2·88-4·03] among boys in the lowest quartile of school performance (Figure S.2 and Table S.5 in the Supplementary Material). When only utilizing the within family variation, the hazard ratio dropped to 3·04 [95% CI 2·38-3·89]. The hazard ratio among girls in the lowest quartile of school performance was 2·59 [95% CI 2·01-3·34] in the benchmark model, and 1·79 [95% CI 1·22-2·63] in the sibling model.

### School performance and causes of young adult death

Utilizing the cause of death register, we estimated the expected number of events for different causes of death across the four quartiles of school performance (Figure 4). Suicide and intentional self-harm were the leading causes of death, constituting 32 percent of the young adult deaths within our sample. Table S.6 in the Supplementary material provides a an overview of the causes included in the cause of death register, with corresponding ICD-10 codes. For the lowest quartiles of school performance there is a considerable share of deaths from poisoning, 38 percent for boys and 36.8 percent for girls. See Table S.7 in the supplementary material for a corresponding table.

**Figure 4:**
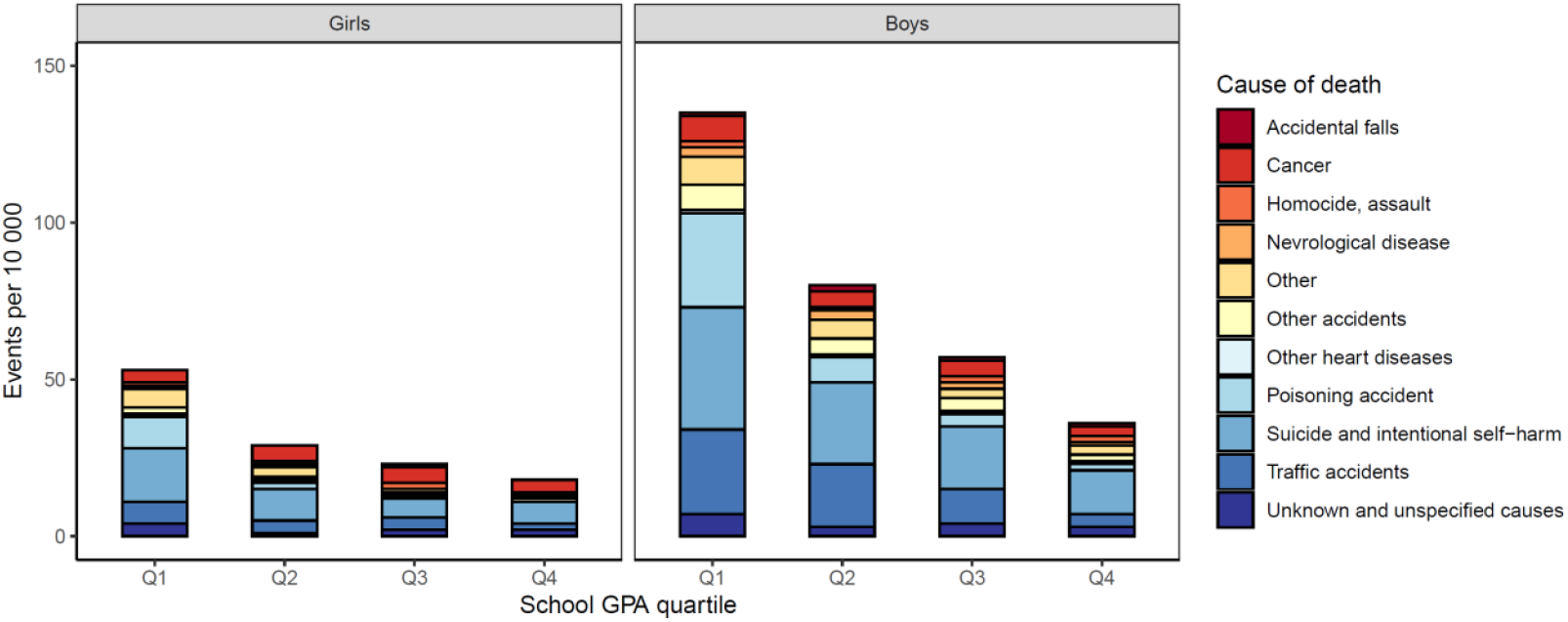
Causes of young adult deaths across sex and quartiles of school performance. The number of deaths were estimated with Cox models (predicted values), separately for each sex, and using dummy variables for each quartile of school performance. The distribution across causes of death were estimated by finding the cause-share with each school performance quartile, then rescaling to the total number of deaths. See table A7 in the supplementary material for a corresponding table.

To examine the relative risk of different causes of death in more details we estimated hazard ratios (HR) for each cause of death among those in the lowest quartile of school performance, relative to the other quartiles (Q2-Q4). The results are displayed in Figure 5. The highest HR ratio was observed for poisoning, with 6·61 [95% CI 5·22-8·37] for boys and 7·89 [95% CI 5·12-12·44] for girls. See Table S.8 in the supplementary material for a corresponding table. Poisoning deaths are primarily related to drug overdoses: For example, in 2018, approximately 80% were from illicit drugs and pharmaceuticals/psychoactive substances (X41-X43), and 10% alcohol poisoning (X45).

**Figure 5:**
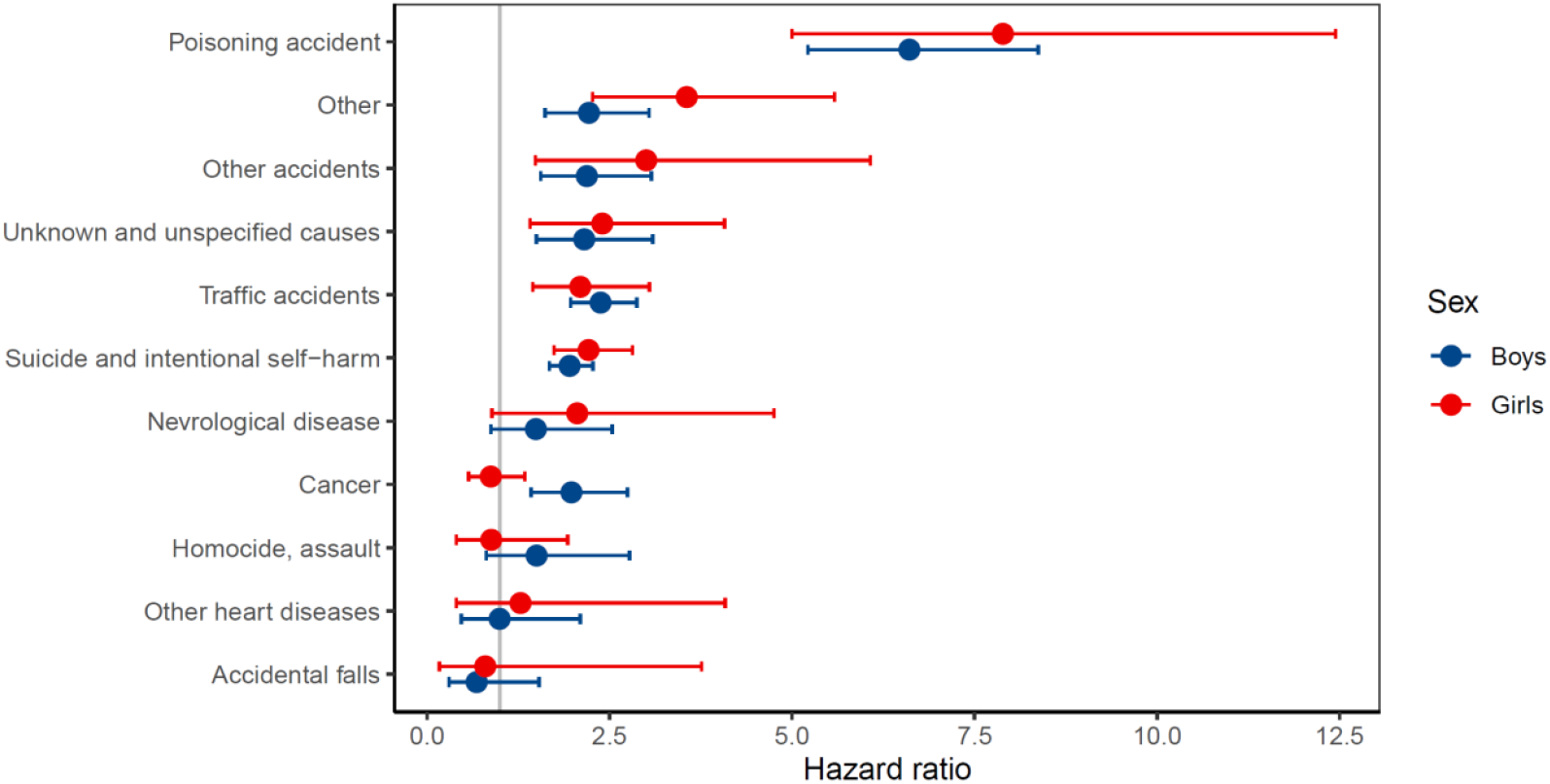
Causes of young adult deaths across sex and quartiles of school performance. Results from Cox regression models run with with young adult death as outcome run separately for each sex and cause of death. See table S.8 in the supplementary material for a corresponding table.

## Discussion

This large population-wide study of 986,573 Norwegian adults has three main findings. First, school performance was a strong predictor of young adult death, substantially stronger than common measures of family socioeconomic background. Second, school performance absorbed the family socioeconomic gradient in young adult death. Third, the primary cause of excessive deaths among those who perform poorly in school was poisoning. To illustrate the magnitude of the association between school performance and young adult death, consider the following examples. The expected reduction in young adult deaths when moving from the 1^st^ to 4^th^ quartile was 100 per 10 000 among boys (from 137 to 37). Or put differently, if everyone in the sample had the same risk as adolescents in the highest quartile, the death rates would have been 52·2% lower (from 0·775% to 0·37%).

This is the first study to show the association between GPA, social background, and causes of young adult death in population-wide individual-level data. Our findings align with studies finding a mortality gradient in educational attainment or IQ.^13, 16, 19^ Especially related is a study from the US finding that the state-level income-mortality gradient could be fully accounted for by the differences in shares of inhabitants with completed high school education.^14^ Documenting the role of school performance, and that it accounts for the family social economic gradient in young adult death, is in itself important because it identifies a group at a particularly high risk. In addition, the following can be inferred from our results about the mechanisms underlying the social gradient: Our study shows that, holding school performance constant, family background does not affect the likelihood of young adult death. Hence, we present evidence suggesting that the traits reflected in school performance are mechanisms through which family socioeconomic status affects health. Which traits do school performance measure? School performance is clearly linked to cognitive ability. But, it also reflects a wider spectrum of executive functioning, such as self-regulation and conscientiousness, which has been shown to be more predictive of future life outcomes than more narrow measures of intelligence.^20^ In summary, our study points to cognitive ability and executive functioning as key mechanisms underlying the family socioeconomic gradient.

In terms of policy implications, this study suggests that policies aimed at reducing young adult death should target adolescents with low school performance or the traits typically associated with low school performance, regardless of the adolescents’ social background. Moreover, death may be seen as the tip of the iceberg of health issues. Hence, this consideration may also be relevant for other health outcomes.

We found a substantially increased risk of young adult death due to poisoning among poor school performers. This finding aligns with recent studies of the mortality development among socially deprived in the US, often referred to as “deaths of despair”. In the US these deaths have contributed to decline in life expectancy in some subgroups of the population.^21^ Closely related to our finding and the literature on “deaths of despair”, a recent study from the US found a decline in life expectancy without a 4-year college degree. Moreover, they show that a large part of the increasing educational differences in life expectancy could be attributed to drug-related deaths. These are often referred to as “preventable deaths”, amenable to behavior change.^8^ Hence, policies aimed at prevention should include social policies next to medical treatment or research.

### Strengths and weaknesses

This study has several strengths, including a large analytic sample covering the whole population, data from high-quality governmental registers, and sibling comparison analyses. Nevertheless, some limitations must be mentioned. First, our results do not allow for causal interpretation. Nevertheless, our findings were robust to two different methods for adjusting to intra-familiar unobservable factors. Hence, environmental and genetic factors shared by siblings appear not to confound the associations. Second, the study is not able to identify which determinants of poor school performance are most important for explaining young adult death. School performance is an outcome caused by a several factors, including cognitive ability, learning environment and personality traits. A deeper understanding of how school performance may affect the likelihood of young adult death would require a more direct measurement of the different factors contributing to young adult death. Third, although we had GPA data on 96% of the relevant population, low school performance and young adult death may be more common among the remaining 4%. Hence, our estimates may underestimate the risks associated with poor performance in lower secondary education.

## Conclusion

School performance accounts for family socioeconomic differences in risk of young adult death. Hence, the family socioeconomic gradient in young adult death can be explained by a combination of cognitive ability and executive functioning. Policy interventions aimed at reducing the social gradient in young adult death should prioritize adolescents with low school performance, regardless of social background. The primary cause of excessive young adult death among those performing poorly in school was drug use. Hence, policies aimed at prevention should be based on social and psychological, rather than biomedical, measures.

## Data Availability

The data for this paper encompasses educational outcomes, income and demographic information for entire cohorts of the Norwegian population. Researchers can access the data by application to the Regional Committees for Medical and Health Research Ethics and the data owners (Statistics Norway and the Norwegian Directorate of Health). The authors cannot share this data with other researchers due to the sensitive nature and potential for identification. However, other researchers should feel free to contact any of the authors if they have questions concerning the data or overlapping research projects.

## Acknowledgements

This work was supported by the Research Council of Norway (grants number 273659 and 300668). This work was partly supported by the Research Council of Norway through its Centres of Excellence funding scheme (grant number 262700). We wish to thank Jonathan Wörn and Anders Skrondal for helpful comments.

## Contributors

The research design was created by B-AR and FAT. B-AR was responsible for data cleaning, data analysis, and reporting. B-AR drafted the article, which was revised by all authors. All authors had final responsibility for the decision to submit for publication. The corresponding author confirms that all authors have seen and approved the final text.

## Supplementary appendix

**Figure S.1:**
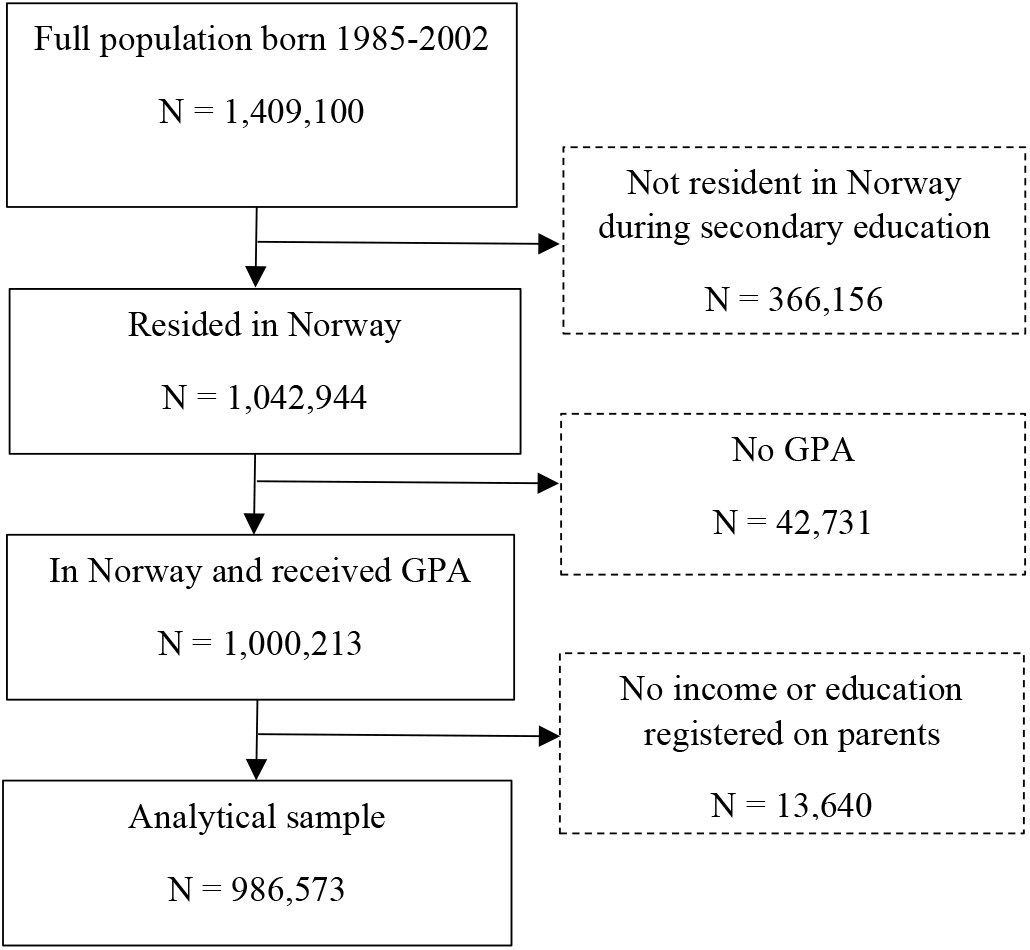
Study sample diagram.

**Table S.1.**
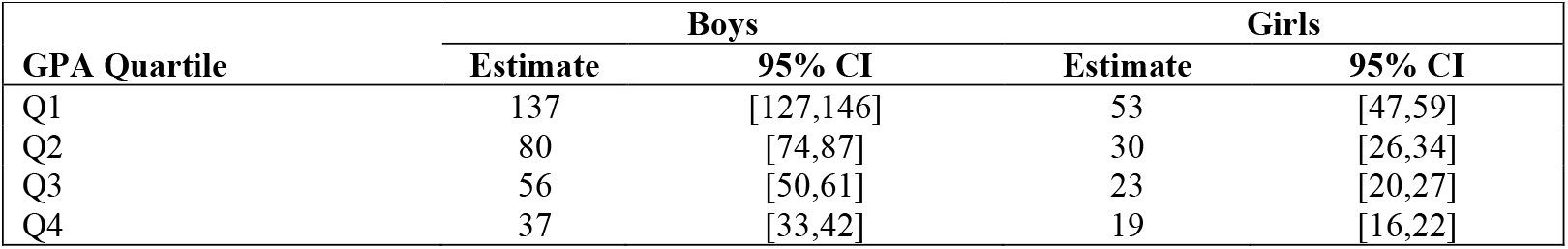
Predicted number of early deaths between age 16 and 30 across sex and quartiles of school performance. Estimated from a Cox regression, separately for each sex, and using dummy variables for each quartile of GPA.

| GPA Quartile | Boys |  | Girls |  |
| --- | --- | --- | --- | --- |
|  | Estimate | 95% CI | Estimate | 95% CI |
| Q1 | 137 | [127,146] | 53 | [47,59] |
| Q2 | 80 | [74,87] | 30 | [26,34] |
| Q3 | 56 | [50,61] | 23 | [20,27] |
| Q4 | 37 | [33,42] | 19 | [16,22] |

**Table S.2.** Predicted number of early deaths between age 16 and 30 across sex and levels of parental education. Estimated from a Cox regression, separately for each sex, and using dummy variables for each level of parental education.

| Parental education | Boys |  | Girls |  |
| --- | --- | --- | --- | --- |
|  | Estimate | 95% CI | Estimate | 95% CI |
| High Uni | 54 | [46,62] | 24 | [19,30] |
| Low Uni | 67 | [62,73] | 28 | [24,32] |
| Secondary | 81 | [76,86] | 33 | [29,36] |
| Primary | 110 | [97,122] | 40 | [32,47] |

**Table S.3.**
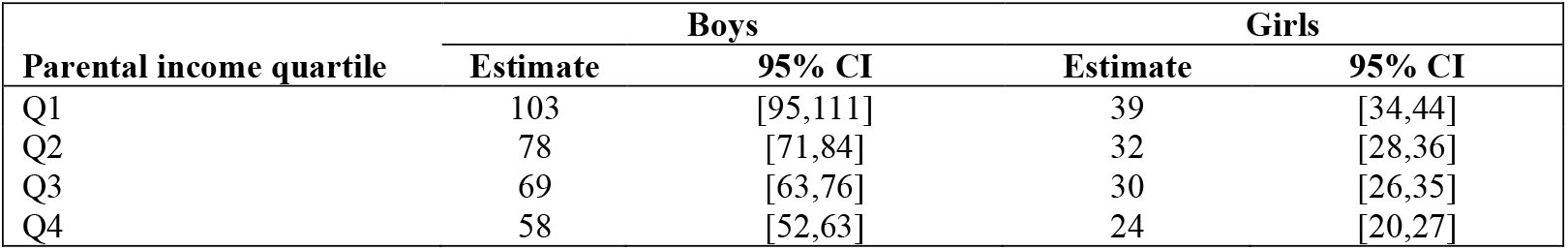
Predicted number of early deaths between age 16 and 30 across sex and quartiles of parental income. Estimated from a Cox regression, separately for each sex, and using dummy variables for each quartile of parental income.

**Table S.4.** Cox regression model: Hazard ratios.

|  | Boys | Girls | Boys | Girls | Boys | Girls | Boys | Girls |
| --- | --- | --- | --- | --- | --- | --- | --- | --- |
| **GPA Q1** | 1.5<br>(1.34, 1.65) | 1.24<br>(1.01, 1.46) |  |  |  |  | 1.49<br>(1.34, 1.64) | 1.24<br>(1.01, 1.47) |
| **GPA Q2** | 2.16<br>(2.01, 2.30) | 1.58<br>(1.36, 1.80) |  |  |  |  | 2.13<br>(1.98, 2.28) | 1.58<br>(1.35, 1.80) |
| **GPA Q3** | 3.66<br>(3.53, 3.80) | 2.82<br>(2.62, 3.02) |  |  |  |  | 3.55<br>(3.41, 3.69) | 2.8<br>(2.59, 3.02) |
| **Parental education: primary** |  |  | 1.25<br>(1.09, 1.42) | 1.16<br>(0.90, 1.41) |  |  | 1.01<br>(0.84, 1.18) | 0.96<br>(0.70, 1.22) |
| **Parental education: secondary** |  |  | 1.5<br>(1.34, 1.66) | 1.35<br>(1.10, 1.59) |  |  | 0.92<br>(0.75, 1.09) | 0.89<br>(0.63, 1.16) |
| **Parental education: low university** |  |  | 2.04<br>(1.86, 2.22) | 1.64<br>(1.35, 1.93) |  |  | 0.99<br>(0.79, 1.19) | 0.87<br>(0.55, 1.19) |
| **Parental income: Q1** |  |  |  |  | 1.2<br>(1.08, 1.33) | 1.29<br>(1.09, 1.49) | 1.05<br>(0.91, 1.18) | 1.17<br>(0.96, 1.38) |
| **Parental income: Q2** |  |  |  |  | 1.35<br>(1.22, 1.47) | 1.35<br>(1.15, 1.55) | 1.05<br>(0.92, 1.18) | 1.12<br>(0.91, 1.33) |
| **Parental income: Q3** |  |  |  |  | 1.79<br>(1.67, 1.91) | 1.63<br>(1.44, 1.83) | 1.25<br>(1.12, 1.38) | 1.24<br>(1.03, 1.46) |
| **Observations** | 505,216 | 481,357 | 505,216 | 481,357 | 505,216 | 481,357 | 505,216 | 481,357 |
| **R2** | 0.001 | 0.0003 | 0.0002 | 0 | 0.0002 | 0.0001 | 0.001 | 0.0003 |
| **Max. Possible R2** | 0.11 | 0.04 | 0.11 | 0.04 | 0.11 | 0.04 | 0.11 | 0.04 |
| **Log Likelihood** | -27,792.88 | -10,688.26 | -28,004.24 | -10,751.16 | -27,992.90 | -10,746.36 | -27,782.80 | -10,685.86 |
| **Wald Test** | 479.51 (df = 3) | 144.36 (df = 3) | 77.93 (df = 3) | 15.35 (df = 3) | 100.97 (df = 3) | 24.41 (df = 3) | 501.65 (df = 9) | 148.90 (df = 9) |
| **LR Test** | 499.75 (df = 3) | 141.10 (df = 3) | 77.02 (df = 3) | 15.31 (df = 3) | 99.71 (df = 3) | 24.90 (df = 3) | 519.90 (df = 9) | 145.91 (df = 9) |
| **Score (Logrank) Test** | 523.23 (df = 3) | 153.50 (df = 3) | 79.30 (df = 3) | 15.48 (df = 3) | 102.79 (df = 3) | 24.72 (df = 3) | 546.18 (df = 9) | 158.15 (df = 9) |

**Note.** \*p<0.1; \*\*p<0.05; \*\*\*p<0.01

**Figure S.2.**
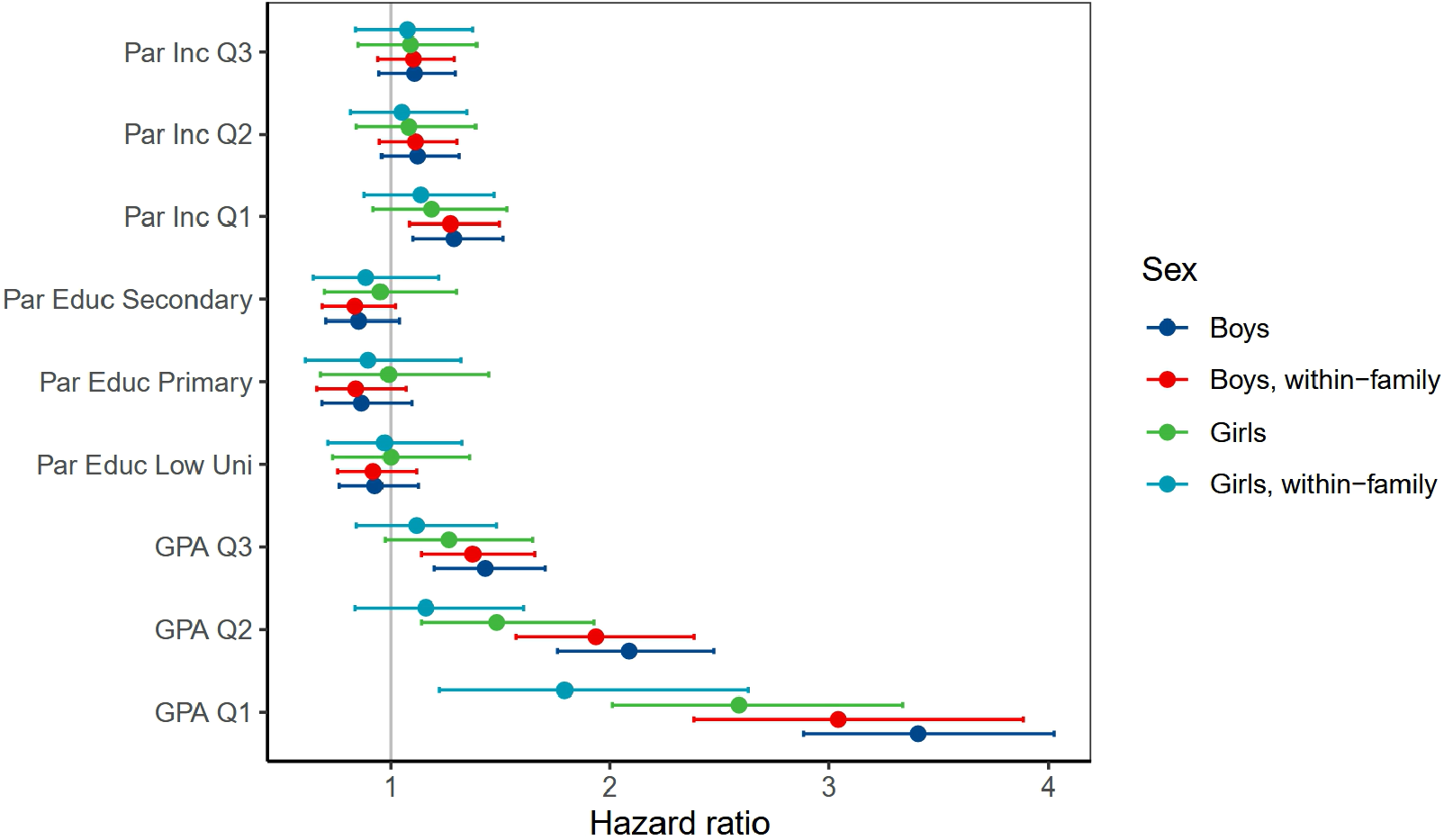
Hazard ratios with and without sibling comparions. Across sex, school performance, parental income and parental education. For comparison, the models were restricted to those with at least one other sibling with a GPA record (N = 740,184). Hazard ratios of early death between age 16 and 30 from a Cox regression, estimated separately for each sex and using dummy variables for each quartile of school performance, quartile of parental income, and highest level of parental education. The within-models were estimated containing both a mother-frailty term and the average school performance (percentile) of the siblings. These models were estimated with the coxph-package in R.

**Table S.5.** Hazard ratios with and without sibling comparions. Across sex, school performance, parental income and parental education. For comparison, the models were restricted to those with at least one other sibling with a GPA record (N = 740,184). Hazard ratios of early death between age 16 and 30 from a Cox regression, estimated separately for each sex and using dummy variables for each quartile of school performance, quartile of parental income, and highest level of parental education. The within-models were estimated containing both a mother-frailty term and the average school performance (percentile) of the siblings. These models were estimated with the coxph-package in R.

| Variable | Boys, HR [95% CI] |  | Girls HR [95% CI] |  |
| --- | --- | --- | --- | --- |
|  | Benchmark model | Within-family | Benchmark model | Within-family |
| GPA Q3 | 1.43[1.2,1.71] | 1.38[1.14,1.66] | 1.27[0.97,1.65] | 1.12[0.84,1.48] |
| GPA Q2 | 2.09[1.76,2.47] | 1.94[1.57,2.38] | 1.48[1.14,1.93] | 1.16[0.84,1.61] |
| GPA Q1 | 3.41[2.88,4.03] | 3.04[2.38,3.89] | 2.59[2.01,3.34] | 1.79[1.22,2.63] |
| Par Educ Low Uni | 0.93[0.77,1.13] | 0.92[0.76,1.12] | 1[0.74,1.36] | 0.97[0.71,1.33] |
| Par Educ Secondary | 0.86[0.7,1.04] | 0.84[0.69,1.02] | 0.95[0.7,1.3] | 0.89[0.65,1.22] |
| Par Educ Primary | 0.87[0.69,1.1] | 0.84[0.66,1.07] | 0.99[0.68,1.45] | 0.9[0.61,1.32] |
| Par Inc Q3 | 1.11[0.95,1.3] | 1.1[0.94,1.29] | 1.09[0.85,1.39] | 1.08[0.84,1.38] |
| Par Inc Q2 | 1.12[0.96,1.31] | 1.11[0.95,1.3] | 1.08[0.84,1.39] | 1.05[0.82,1.35] |
| Par Inc Q1 | 1.29[1.1,1.51] | 1.27[1.09,1.5] | 1.19[0.92,1.53] | 1.14[0.88,1.47] |
| Sibling GPA percentile | - | 1[0.99,1] | - | 0.99[0.99,1] |
| N | 378869 | 378869 | 360097 | 360097 |

**Table S.6.**
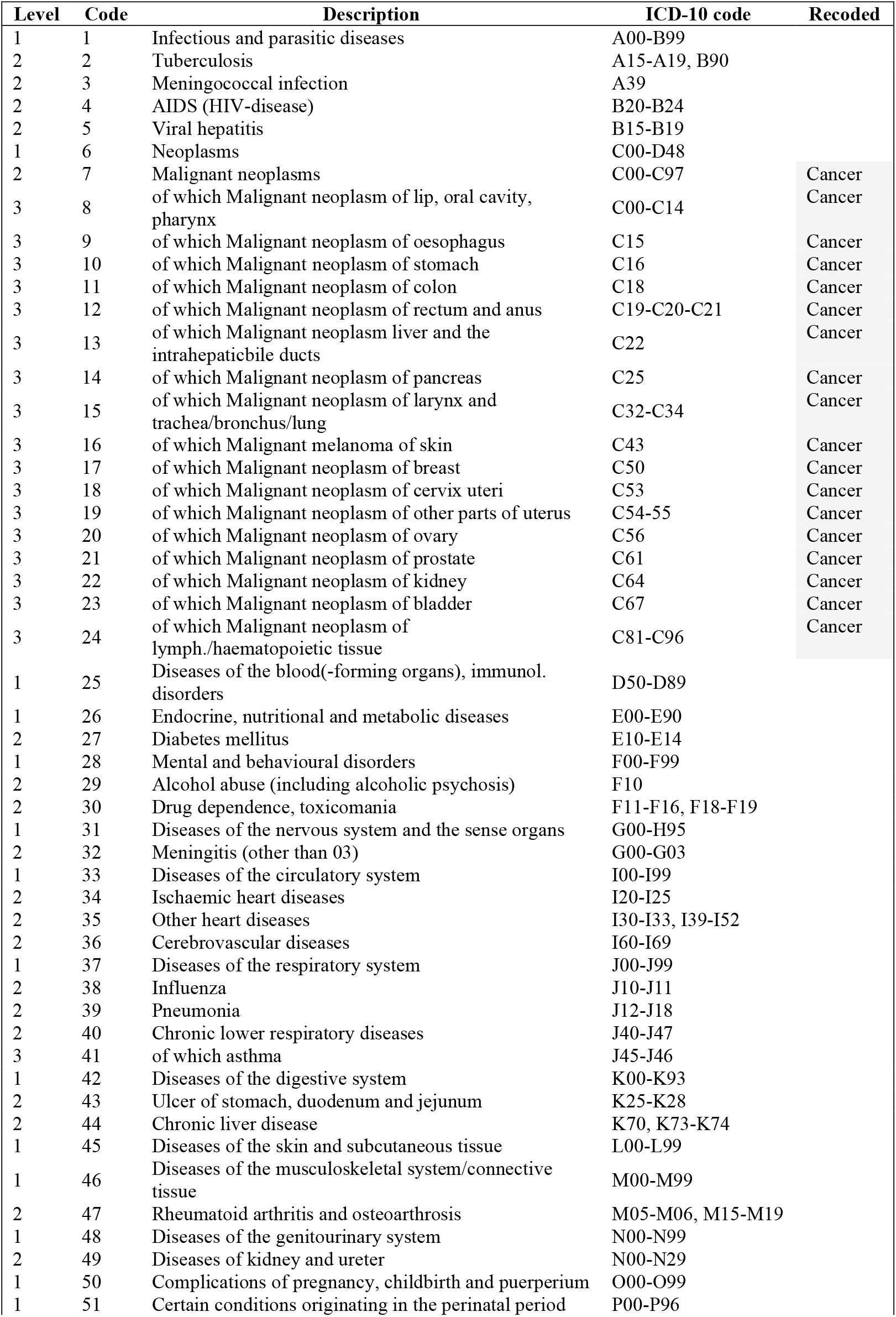
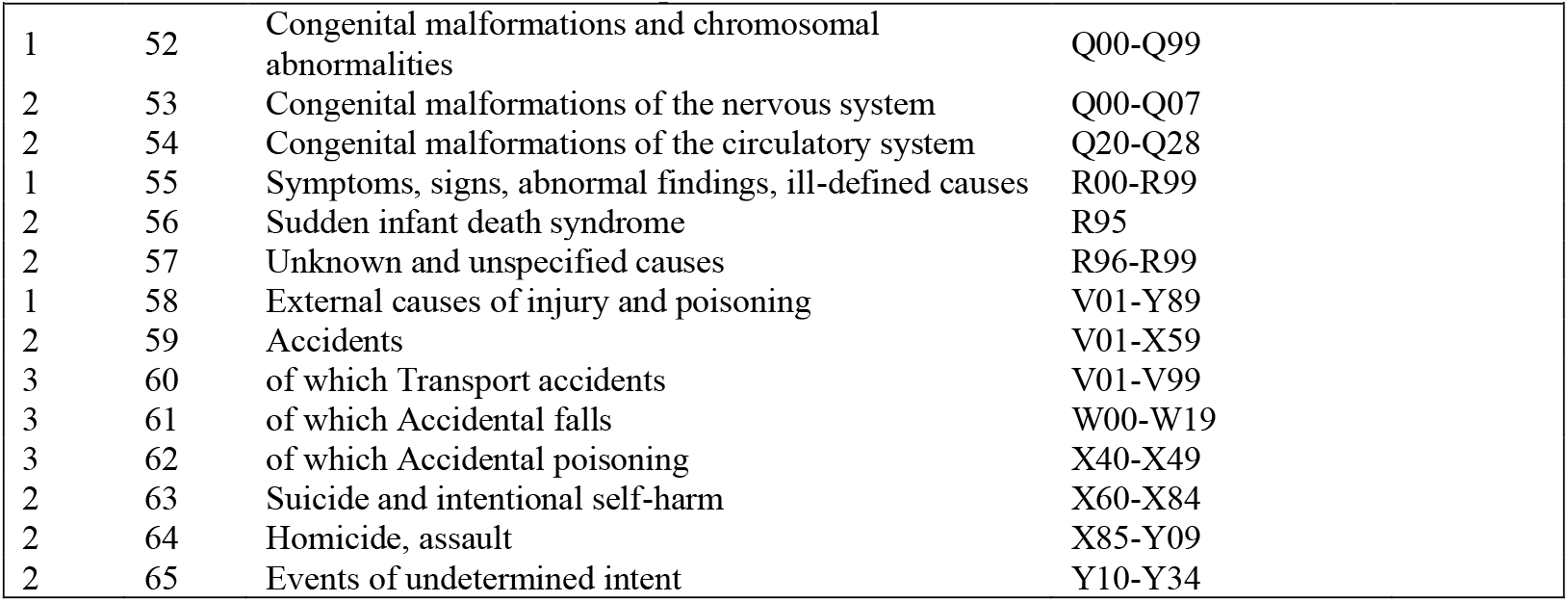
EU Shortlist Causes of Death.

**Table S.7.** Estimated number of early deaths per 10 000 between age 16 and 30 by causes of early death, across sex and quartiles of school performance. Estimated from a Cox regression, separately for each sex, and using dummy variables for each quartile of GPA.

| Cause of death | Girls |  |  |  | Boys |  |  |  |
| --- | --- | --- | --- | --- | --- | --- | --- | --- |
|  | Q1 | Q2 | Q3 | Q4 | Q1 | Q2 | Q3 | Q4 |
| Accidental falls | 0.303 | 0.145 | 0.716 | 0.284 | 1.012 | 1.913 | 1.087 | 1.485 |
| Cancer | 4.097 | 5.491 | 5.01 | 3.687 | 8.388 | 4.918 | 4.756 | 3.105 |
| Homocide, assault | 1.214 | 1.012 | 1.861 | 1.276 | 2.169 | 1.093 | 1.63 | 1.62 |
| Nevrological disease | 1.366 | 1.301 | 0.286 | 0.425 | 2.892 | 3.415 | 1.766 | 0.675 |
| Other | 6.07 | 2.89 | 1.002 | 1.276 | 9.4 | 6.421 | 3.397 | 2.97 |
| Other accidents | 2.276 | 1.012 | 0.573 | 0.709 | 8.098 | 4.781 | 4.076 | 2.295 |
| Other heart diseases | 0.607 | 0.723 | 0.286 | 0.425 | 1.302 | 1.23 | 1.495 | 1.215 |
| Poisoning accident | 9.712 | 2.457 | 1.002 | 0.284 | 30.369 | 7.787 | 3.669 | 2.43 |
| Suicide and intentional self-harm | 16.996 | 9.827 | 6.442 | 6.949 | 38.757 | 26.093 | 19.566 | 14.174 |
| Traffic accidents | 6.98 | 4.046 | 4.008 | 1.985 | 27.043 | 19.536 | 10.598 | 4.185 |
| Unknown and unspecified causes | 3.642 | 0.867 | 2.147 | 1.56 | 7.086 | 3.142 | 3.669 | 3.105 |

**Table S.8.** Causes of early death between age 16 and 30 across sex and quartiles of school performance: Hazard ratios for those with poor school performance. Estimated from a Cox regression, separately for each sex, and using a dummy variables for the first quartile of GPA performance.

| Cause of death | Girls |  | Boys |  |
| --- | --- | --- | --- | --- |
|  | Hazard ratio | 95% CI | Hazard ratio | 95% CI |
| Accidental falls | 0.8 | [0.17,3.76] | 0.68 | [0.3,1.54] |
| Cancer | 0.87 | [0.57,1.34] | 1.98 | [1.43,2.75] |
| Homocide, assault | 0.88 | [0.4,1.93] | 1.5 | [0.81,2.77] |
| Nevrological disease | 2.06 | [0.89,4.75] | 1.49 | [0.88,2.54] |
| Other | 3.56 | [2.27,5.58] | 2.22 | [1.62,3.04] |
| Other accidents | 3.00 | [1.49,6.08] | 2.19 | [1.56,3.08] |
| Other heart diseases | 1.28 | [0.4,4.09] | 1.00 | [0.47,2.1] |
| Poisoning accident | 7.89 | [5,12.44] | 6.61 | [5.22,8.37] |
| Suicide and intentional self-harm | 2.21 | [1.74,2.82] | 1.95 | [1.68,2.27] |
| Traffic accidents | 2.1 | [1.45,3.05] | 2.38 | [1.97,2.87] |
| Unknown and unspecified causes | 2.4 | [1.41,4.08] | 2.15 | [1.5,3.09] |

